# BEYOND BONES - THE RELEVANCE OF VARIANTS OF CONNECTIVE TISSUE (HYPERMOBILITY) TO FIBROMYALGIA, ME/CFS AND CONTROVERSIES SURROUNDING DIAGNOSTIC CLASSIFICATION: AN OBSERVATIONAL STUDY

**DOI:** 10.1101/2020.02.21.20025072

**Authors:** Jessica A Eccles, Beth Thompson, Kristy Themelis, Marisa Amato, Robyn Stocks, Amy Pound, Anna-Marie Jones, Zdenka Cipinova, Lorraine Shah-Goodwin, Jean Timeyin, Charlotte R Thompson, Thomas Batty, Neil A Harrison, Hugo D Critchley, Kevin A Davies

## Abstract

**Objectives:** To understand the relevance of symptomatic hypermobility and related connective tissue variants to the expression of symptoms in Fibromyalgia and Myalgic Encephalomyelitis/Chronic Fatigue Syndrome (ME/CFS). The study further tested if specific subfactors within the diagnostic classification of hypermobility predict clinical presentations.

**Design:** We report part of a larger case-control study exploring mechanisms of chronic pain and fatigue in Fibromyalgia and ME/CFS (https://doi.org/10.1186/ISRCTN78820481)

**Setting:** an NHS Clinical Research Facility.

**Participants:** A subsample of 87 participants were assessed for symptomatic hypermobility by a trained clinician: 63 presented with a clinical diagnosis of either Fibromyalgia and or ME/CFS confirmed at screening; 24 participants were confirmed as healthy controls.

**Main outcome measures:** 1) Brighton Criteria for joint hypermobility syndrome and 2017 hEDS diagnostic criteria. 2) ACR 2010 Diagnostic Criteria for Fibromylagia and Canadian and Fukada diagnostic criteria for ME/CFS. 3) Self report measures of subjective pain, fatigue and interoceptive sensibility.

**Results:** Twenty of the 63 patients (32%) presented with a clinical diagnosis of Fibromyalgia; 24 (38%) with a clinical diagnosis of ME/CFS and 19 (30%) with dual diagnoses of fibromyalgia and ME/CFS. After evaluation using clinical research tools, 56 patients (89%) met ACR diagnostic criteria for fibromyalgia; 59 (94%) Canadian Criteria for ME/CFS; and 61 (97%) Fukada Criteria for ME/CFS. After research evaluation 52 patients (85%) in fact met diagnostic criteria for Fibromyalgia and ME/CFS on all three sets of tools (ACR, Canadian, Fukada). In addition, 51 patients (81%) and 9 (37.5%) healthy controls met Brighton Criteria for joint hypermobility syndrome and 11 (18%) and 2 (8%) of patients and controls respectively, on the 2017 hEDS criteria. Of these patient participants with symptomatic hypermobility only 12 (23.5%) had received a prior diagnosis of hypermobility.

Across all participants meeting Brighton Criteria, 13 (22%) also endorsed a hEDS diagnosis. Membership of the patient group was predicted by meeting the Brighton Criteria for joint hypermobility syndrome (p=<0.001, OR 7.08, 95%CI 2.50 – 20.00), but not by meeting the hEDS criteria. The historical, rather than current Beighton score correlated with; 1) total pain reported on the McGill Pain Questionnaire (short form), (r= 0.25, n= 73, p=0.03); 2) Widespread Pain Index (derived from ACR diagnostic criteria) (r=0.26, n= 86, p=0.01); 3) ACR symptom severity (r=0.27, n=85, p=0.01); 4) Fatigue Impact (r=0.29, n=56, p=0.28); and 5) interoceptive sensibility (r=0.30, n=56, p=0.02).

**Conclusions:** Symptomatic joint hypermobility is relevant to symptoms and diagnosis in Fibromyalgia and ME/CFS. These conditions are poorly understood yet have a considerable impact on quality of life. Further work is needed to determine the prevalence of hEDS within the general population and define the critical clinical dimensions within symptomatic hypermobility. It is important to note the high rates of mis/underdiagnosis of symptomatic hypermobility in this group. Moreover, we need to clarify the role of variant connective tissue in dysautonomic and inflammatory mechanisms implicated in the expression of pain and fatigue in fibromyalgia and ME/CFS. Our observations have implications for diagnosis and treatment targets.

**Study registration:** ISCRTN78820481

## INTRODUCTION

Fibromyalgia and Myalgic Encephalomyelitis/Chronic Fatigue Syndrome (ME/CFS) have overlapping symptoms. Fibromyalgia is a common, poorly understood, complex musculo-skeletal disorder that affects around 5% of the UK population^1^. It is polysymptomatic yet marked primarily by chronic widespread pain, typically believed to be non-inflammatory in nature. In addition, patients frequently report extra-musculoskeletal symptoms, including both physical and emotional fatigue with subjective cognitive dysfunction (‘fibrofog’^2 3^). For quality of life, patients with fibromyalgia score significantly lower than the general population^2^, or patients with other musculo-skeletal disorders such as rheumatoid arthritis, osteoarthritis and SLE^4^, on all-eight health status domains. Pain, fatigue, subjective cognitive impairment and psychological disturbance (e.g. depression and anxiety)^5^ have greatest impact on quality of life. Thus, pain and fatigue are identified as among the most disabling symptoms associated with fibromyalgia. Relatedly, pain is also frequently reported in ME/CFS^6 7^. Complexity to understanding their origins may have contributed at times to consideration of both fibromyalgia and ME/CFS as functional somatic, medically-unexplained, or somatisation disorders. The presence of shared symptoms in brain and body has also fuelled debate as to whether fibromyalgia and ME/CFS are manifestations of the same spectrum disorder or separate clinical entities^8 9^. Although the dominant symptom of fibromyalgia is pain and ME/CFS is disabling fatigue or post-exertional malaise, patients with either diagnosis typically experience pain (including hyperalgesia and central sensitisation), fatigue, sleep problems and cognitive difficulties^9^. Altered inflammatory markers^10 11^ and dysautonomia^12-14^ are reported in both fibromyalgia and ME/CFS, calling into question their perceived status as ‘functional’ disorders. Moreover, it is increasingly recognized that fibromyalgia^15^ and ME/CFS^5 16 17^ are associated with joint hypermobility ^18-21^, an expression of variant connective tissue. In this context, most patients (82.6%) with Ehlers Danlos Syndrome –Hypermobility Type (EDS-HT) /Joint Hypermobility Syndrome patients (JHS) meet criteria for ME/CFS, compared to only 8.4% of controls^22^. Beighton (hypermobility) scores are higher in children with a ME/CFS diagnosis than gender-matched controls^23^. Similarly, criteria for joint hypermobility is reportedly met by 64.2% women with fibromyalgia compared to only 22% of controls^24^. Clinicians may diagnose hypermobility as fibromyalgia, since widespread pain is a key feature of both conditions^25^.

Joint hypermobility (including Ehlers-Danlos Syndrome(s)) ^18-21^ is common^26 27^ and associated with many extra-articular features^28^. Internationally, there is little agreement on the precise definition of measurement and hypermobility, nor the point at which it constitutes a disorder. In 2017, a consensus group sought to re-classify Ehlers Danlos Syndrome-Hypermobility Type (EDS-HT). As a consequence, the terms hEDS (hypermobile EDS) and Hypermobility Spectrum Disorder (HSD) now refer to symptomatic joint hypermobility^29^. An earlier term, Joint Hypermobility Syndrome (diagnosed by Revised Brighton Criteria)^30^ was made redundant and was previously deemed indistinquishable from EDS-HT^31^. However, one important distinction is that the Revised Brighton Criteria took into account historical joint hypermobility, whereas the newer criteria for hEDS rely on age and gender specific hypermobility cut-offs. A broader concept of a Hypermobility Spectrum Disorder (HSD) was introduced to describe the continuum that encompasses patients with symptomatic joint hypermobility through to people who do not meet full criteria for hEDS or related syndromes associated with joint hypermobility. However, this notion of a spectrum also remains controversial. Since there are no genetic tests available for hEDS, a clinical diagnostic checklist is available and consistent with the 2017 consensus criteria^29^. Further research is required to validate clinical diagnostic criteria and international diagnostic codes, which presently are not consistent with newer nomenclature. In adults with hEDS/HSD, symptoms (including pain, fatigue, dysautonomic symptoms, co-ordination, attention/concentration deficits and quality of life in general) can be also classified according to severity, reflecting neither the old or new criteria for hEDS^32^. Debate regarding diagnostic classification has intensified following a report on the prevalence of EDS and JHS in primary care that suggests these diagnoses cannot be considered rare^33^. Moreover, symptomatic hypermobility has considerable impact on quality of life, at least comparable or greater than those with other rheumatological diseases.^34-45^

The morbidity of hEDS/HSD to patients is most commonly expressed as complaints to clinicians of chronic musculoskeletal pain, headache and fatigue. These symptoms impair physical activity and quality of life^34-45^, common to both FIbromyalgia and ME/CFS. In rheumatology clinics, symptomatic hypermobility is estimated to contribute to nearly half of all outpatient appointments, but is only recognised 1 out of 19 times^46^. Under-diagnosis of hEDS/HDS may exacerbate potentially disabling symptoms complications^47^ and comorbidities, including autonomic dysfunction^14 48^, fibromyalgia^24^ and myalgic encephalomyelitis/chronic fatigue syndrome (ME/CFS)^49^. Enhanced recognition of symptomatic hypermobility is essential to improve function and quality of life in a patient group, whose symptoms are typically poorly understood and frequently overlooked^46^. Accurate phenotyping of hEDS/HDS is needed for research, but it is also critical to effective multi-disciplinary clinical management, from provision of accurate prognostic advice and identification of co-morbidity, to the offer of multi-professional support and delivery of effective therapies. The relevance extends beyond rheumatology to other clinical contexts; e.g. cardiology, paediatrics, neurology, gastroenterology, pain management and mental health settings.

### Aims of the study

The objective of this study was to understand the relevance of symptomatic hypermobility and related connective tissue variants to the expression of symptoms in Fibromyalgia ME/CFS. The study systematically tests associations between the diagnostic criteria for pain and fatigue symptom expression and further examines if specific sub factors within the diagnostic classifications of hypermobility predict clinical presentations.

## METHODS

This study is part of larger study investigating mechanisms of chronic pain and fatigue in fibromyalgia and ME/CFS (https://doi.org/10.1186/ISRCTN78820481) in adults. This paper focuses on the relevance of symptomatic hypermobility. All participants (patient participants and healthy controls) provided written informed consent and were compensated for their participation.

### Patient and public involvement

The study idea was formed after patients and prior research participants discussed their lived experience of fibromyalgia and ME/CFS with the clinicians involved in the initial grant application (JAE, NAH, KAD, HDC). A formal Patient and Public Involvement panel (Brighton and Sussex University Hospitals Trust JAFFA Panel) reviewed the study after funding was obtained and advised on the study protocol and design, strategies for recruitment reflecting their priorities, experiences and preferences. Patients and public assessed the burden of testing and the time required to participate in the study. Patients and public were involved in the promotion of the study through local support groups for patients living with fibromyalgia and ME/CFS. Patients and public will be involved in supporting the dissemination of the study findings to participants and wider relevant communities

Patient participants with a clinical diagnosis of either fibromyalgia and/or ME/CFS were invited to be screened for the study. To be included in the study patients were required to either meet the American College of Rheumatology (ACR 2010)^50^ diagnostic criteria for FM and/or both the Canadian^51^ and Fukada (CDC)^52^ Criteria for ME/CFS as assessed at screening. Patients were recruited by advertisement from local support organisations for Fibromyalgia and ME/CFS, local rheumatology clinics and via social media and bulletin boards. Inclusion criteria for healthy controls were as follows: A score no more than 3/10 on a pain visual analogue scale, a score of less than 36 on the Fatigue Severity Scale^53^ and not meeting either the criteria for American ACR diagnostic criteria for fibromyalgia, or the Canadian and Fukada (CDC) Criteria for ME/CFS and be free of major psychiatric or neurological illness. Healthy controls were also recruited via social media and bulletin boards. Neither patients nor controls were recruited/ selected or screened on the basis of hypermobility.

In total, 24 healthy controls were assessed alongside 63 patient participants. Where possible controls were matched to age and gender of patient participants.

All participants who responded to advertisements were provided with more information. If they were willing to continue, there were screened via telephone or via anonymized online questionnaires for eligibility. Consenting participants completed a series of baseline questionnaires measuring subjective pain (McGill Pain short form Questionnaire^54^); fatigue (Fatigue Impact^55^, Modified Fatigue Impact Scale^56^); brain-fog (Mental Clutter Scale^57^) and interoceptive sensibility^58^ (Porges Body Perception Questionnaire^59^). Demographic data was also collected and participants were assessed for presence of symptomatic hypermobility and variants of connective tissue by trained clinicians using the revised Brighton Criteria and the 2017 hEDS classification criteria^29 46^. It was not possible to determine the last item in criterion 2A (Aortic root dilatation with Z-score >+2) as participants did not undergo echocardiography as part of the study protocol.

### Data analysis and statistical methods

Each symptomatic hypermobility diagnostic classification (revised Brighton Criteria for JHS; hEDS 2017 consensus criteria) were entered as binary variables. Further binary variables explored categorical elements of the different criteria. The Beighton Scale and total number of major and minor criteria endorsed were expressed as scale data. Differences between groups in age and assessment outcomes were assessed via independent sample t tests. Differences in sex and associations between groups were assessed via chi square test. Binary logistic regression was used to derive prediction models for catergorical variables, their odds ratios and their confidence intervals where appropriate. Univariate analyses (General Linear Model) were used to determine interactions. Spearman correlations were used to explore correlations with Beighton score. Hypothesis tests were considered statistically significant for p-values < 0.05 and all analyses were carried out using SPSS version 25 for Mac.

## Results

### Patient characteristics and diagnostic groups

Twenty-four healthy controls (9 male) and patients (12 male) did not differ significantly in age. Mean (SEM) age of all healthy controls (39.3 years, (2.7)) was not significantly different to the whole patient group (45.4, 1.9). There were no significant differences in sex.

Among the 63 patients, 20 (32%) presented with a clinical diagnosis of fibromyalgia; 24 (38%) presented with a clinical diagnosis of ME/CFS; and 19 (30%) presented with dual diagnoses of fibromyalgia and ME/CFS

On research diagnostic evaluation, 56 (89%) met ACR diagnostic criteria for fibromyalgia, 59 (94%) met Canadian Criteria for ME/CFS and 61 (97%) met Fukada Criteria for ME/CFS. Strikingly,. fifty-two patients (85%) met all three diagnostic criteria for fibromyalgia and ME/CFS (ACR, Canadian, Fukada). Nine (37.5%) healthy controls met Brighton Criteria, compared to 51 (81%) of the patient group. Two (8%) of healthy controls met 2017 hEDS criteria, compared to 11 (18%) of the patients. Across all participants meeting Brighton Criteria,13 (22%) endorsed a hEDS diagnosis. Of those assessed as having symptomatic hypermobility only 12 (23.5%) had received a diagnosis of hypermobility prior to evaluation in this study.

### Relevance of symptomatic hypermobility to diagnosis

#### Beighton score

Across all participants there were no associations between current (i.e. present) Beighton score and baseline pain, fatigue and interoceptive questionnaires. Historical Beighton score (i.e ever being able to perform the relevant manoeuvre) correlated positively with total pain on McGill Pain short form Questionnaire^54^ (r= 0.25, n= 73, p=0.03), Widespread Pain Index (derived from ACR diagnostic criteria) (r=0.26, n= 86, p=0.01), ACR somatic symptom severity (r=0.27, n=85, p=0.01), Fatigue Impact(r=0.29, n=56, p=0.28), interoceptive sensibility (r=0.30, n=56, p=0.02) (all spearman rank correlations).

There was a significant interaction of being a patient with fibromyalgia and/or ME/CFS compared to being a control on the relationship between historical Beighton score and McGill Pain Score (F=5.50, p=0.022); Widespread Pain Index (F=36.87, p<0.001); ACR somatic symptom severity (F=59.74, p=<0.001); Fatigue Severity Scale (F=35.29, p=<0.001); Fatigue Impact (F=27.37, =<0.001); Modified Fatigue Impact Scale (F=37.01, p=<0.001); Mental Clutter Scale (F=15.72, p=<0.001)(Univariate General Linear Model). This means that the relationship symptoms and being a patient and control was significantly different according to hypermobility score

#### Brighton Criteria for Joint Hypermobility Syndrome

##### 1. Mixed Fibromyalgia/ME/CFS

Meeting Brighton Criteria significantly predicted membership of the patient group (OR 7.08, p=<0.001, 95%CI 2.50 – 20.00), and meeting both major criteria for the Brighton Criteria was significantly associated with being a patient *(□* ^*2*^ *=9*.*26, p= 0*.*022)*

Easy bruising was significantly associated with being a patient rather than a control (□^*2*^ *4*.*10, p= 0*.*043)*. No other minor criteria were significantly predictive of group membership. There was a significant interaction of joint hypermobility syndrome on the relationship between interoceptive sensibility and group membership (F=8.75, p=0.005)

##### 2. Fibromyalgia

Endorsing both major criteria of Brighton Criteria was significantly associated with an ACR fibromyalgia diagnosis(□^2^ 6.53, p=0.01). ACR Fibromyalgia was associated with easy bruising, (□^2^ 7.58, p= 0.006). No other minor criteria were significantly predictive of fibromyalgia

##### 3. ME/CFS

Endorsing both major criteria of Brighton Criteria was significantly associated with fulfilling Canadian criteria for ME/CFS (□^2^ 7.64, p= 0.006), as was endorsing Fukada Criteria (□^2^ 10.50, p= 0.001). Easy bruising (□^2^ 4.22, p=0.040) was associated with both Canadian criteria for ME/CFS and Fukada Criteria (□^2^ 5.71 p= 0.017). No other minor criteria were significantly predictive of ME/CFS

##### 4. Meeting all 3 diagnostic criteria (ACR, Canadian, Fukada)

Meeting Brighton Criteria for Joint Hypermobilty Syndrome predicted meeting all three patient diagnostic criteria (p<0.001, OR 8.57 95%CI 3.02 – 24.05)

#### hEDS 2017 Diagnositic Criteria

##### 1. Mixed Fibromyalgia and or//ME/CFS

A diagnosis of hEDS did not predict presence of Fibromaylgia and or/ME/CFS, nor did individual elements of the diagnostic criteria

##### 2. Fibromyalgia

There were no associations between ACR definition of fibromyalgia and hEDS diagnosis or elements of the diagnostic criteria

##### 3. ME/CFS

Endorsing hEDS criterion 1 (age/sex adjusted Beighton score) significantly predicted presence of ME/CFS as defined by Canadian criteria (p=0.045, OR 3.07 95% CI (1.01 – 9.28)), and the Fukada Criteria (p=0.031 OR 3.70 95% CI 1.12 – 12.18) No other elements of the hEDS diagnostic criteria were significantly predictive

##### 4. Meeting all 3 diagnostic criteria (ACR, Canadian, Fukada)

A diagnosis of hEDS did not significantly predict meeting all three criteria. Neither did any diagnostic element

## DISCUSSION

The strength of this research study is that it is the first clinical evaluation to our knowledge, that incorporates and directly compares Brighton (Joint Hypermobility Syndrome) and hEDS criteria. This study demonstrates the overlapping nature of Fibromyalgia and ME/CFS within current diagnostic criteria, which remains controversial ^8 9^. We show the contribution of symptomatic joint hypermobility and variants of connective tissue disorder to both conditions and to associated level of symptoms. Fibromyalgia and ME/CFS are both associated with inflammatory abnormalities^10 11^ and dysautonomia^12-14^ (e.g. orthostatic intolerance, postural orthostatic tachycardia syndrome and orthostatic hypotension), which is also commonly observed in symptomatic hypermobility ^14 48^. In fact, autonomic dysfunction is proposed to mediate disabling symptoms in fibromyalgia and has an established association with variants of connective tissue and heightened interoceptive sensibility ^60^. Interoceptive sensibility is subjective measurement of the sense relating to one’s own internal bodily sensations ^58^ While it is clear that there are associations between symptomatic hypermobility and both fibromyalgia and ME/CFS, the relationships between these conditions remain poorly understood. Ongoing research seeks to extend our observations reported here to determine how symptomatic hypermobility relates to autonomic and inflammatory induced changes in pain and fatigue in fibromyalgia and ME/CFS. Limitations of the present study include relatively small control group and difficulty in discriminating between pain-predominant and fatigue-predominant disease.

This paper adds further recognition to the high rates of hypermobility in fibromyalgia and ME/CFS. However, this is the first study to use multiple complementary assessments of joint hypermobility and variant connective tissue to explore these relationships. A rich debate has been stimulated by a recent paper reporting the prevalence of EDS and JHS^33^. While other forms of EDS are likely rare, the present study suggests hEDS is common among patients with pain and fatigue. Our control group is too small to make more than anecdotal statements regarding hEDS prevalence. However, approximately 1 in 5 of those meeting Brighton Criteria for joint hypermobility syndrome might endorse a diagnosis of hEDS and many patients appear to remain undiagnosed.

The classification of symptomatic joint hypermobility and variants of connective tissue is an evolving area^29 31 61 62^. Importantly, this study reveals that distinct aspects of the diagnostic criteria for symptomatic hypermobility predict symptom severity in pain and fatigue conditions. From these data, it would appear that the Brighton Criteria predict Fibromyalgia and ME/CFS diagnosis and symptomatology Interestingly, the Brighton Criteria make use of the historical account of joint laxity, which was highly associated with symptom severity, compared to current joint laxity, which is not included within the hEDS 2017 criteria (Criteria 1).

This relatively small study highlights the need for better recognition and accurate diagnosis in these poorly understood conditions. Fibromyalgia patients may wait on average almost a year after experiencing symptoms before presenting to a physician, and a diagnosis of fibromyalgia may take around 2.3 years, presenting to 3.7 different physicians, to be established ^63^. Once confirmed a fibromyalgia diagnosis may have mixed benefits for patients^64^. Similar delays to diagnosis are observed for ME/CFS^65^. The impact on quality of life in Fibromyalgia and ME/CFS is severe^66 67^ Greater awareness among the scientific and medical community about the overlapping nature of these conditions and their system-wide symptomatology will improve the appropriate diagnosis and multi-disciplianry treatment for patients ^68^. Studies underway will determine the wider role of variant connective tissue (of which joint hypermobility may be but a single manifestation) in relationship to autonomic and inflammatory induced changes in pain and fatigue in Fibromyalgia and ME/CFS (https://doi.org/10.1186/ISRCTN78820481) including interoceptive and neural mechanisms and the role of the transcriptome.

**Figure 1.**
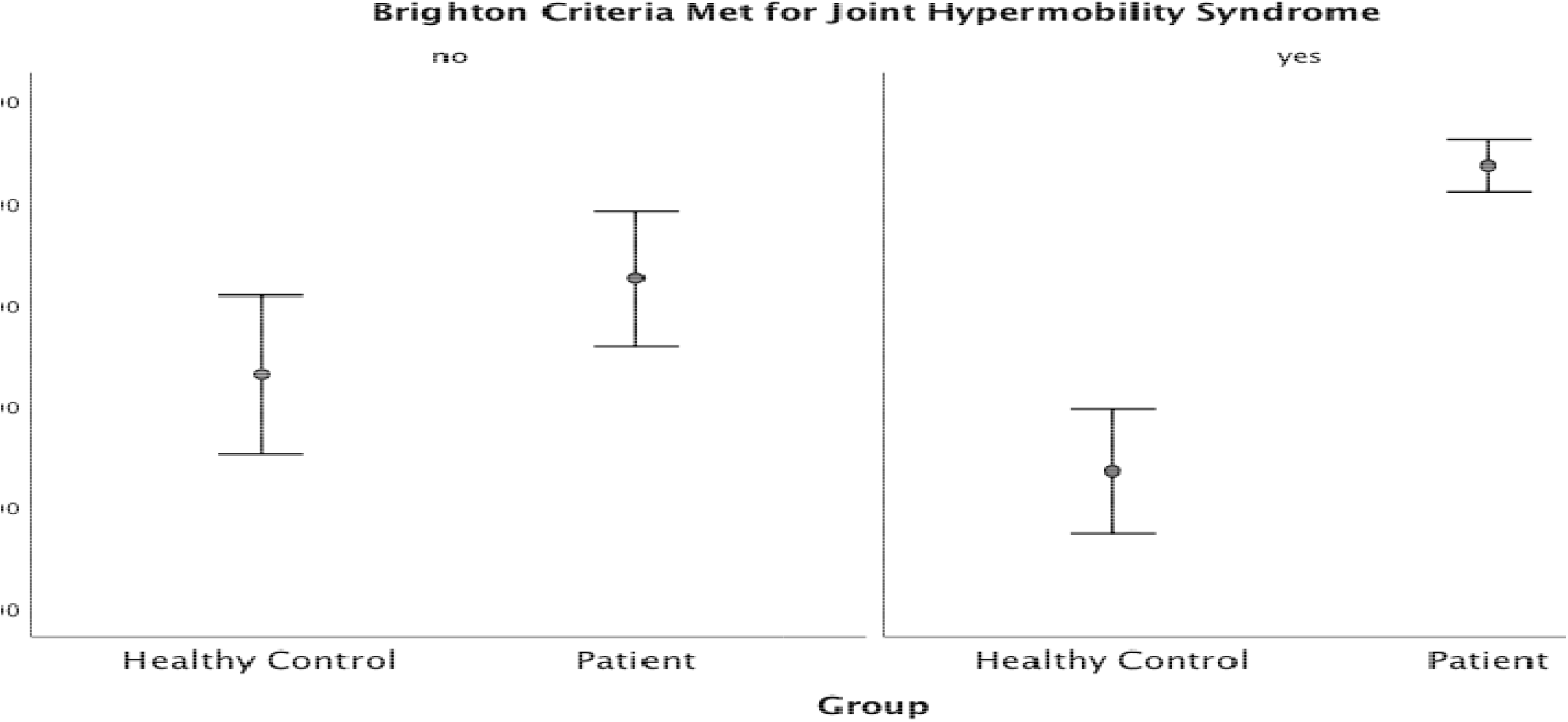
Interaction of joint hypermobility syndrome on the relationship between interoceptive sensibility and Fibromyalgia and or ME/CFS

## Data Availability

Deindentified participant data is available upon reasonable request from the corresponding author (ORCID ID https://orcid.org/0000-0002-0062-1216).

## CONTRIBUTORSHIP AND GUARANTOR INFORMATION

Jessica Eccles, Neil Harrison, Hugo Critchley and Kevin Davies designed the study. Jessica Eccles, Kristy Themelis, Marisa Amato, Zdenka Cipinova, Lorraine Shah-Goodwin, Jean Timeyin, Thomas Batty and Charlotte Thompson collected data for the study. Beth Thompson, Robyn Stocks and Amy Pound processed data for the study. Jessica Eccles analysed data for the study with the assistance of Anna – Marie Jones. Jessica Eccles, Kevin Davies, Hugo Critchley, Beth Thompson, Anna Marie Jones and Neil Harrison drafted the manuscript. All authors approved the manuscript. Jessica Eccles acts a guarantor of the study. The guarantor accepts full responsibility for the work and/or the conduct of the study, had access to the data, and controlled the decision to publish. The corresponding author attests that all listed authors meet authorship criteria and that no others meeting the criteria have been omitted.

## ACKNOWLEDGEMENTS

We wish to acknowledge the invaluable practical assistance of Valentina Toska and Mel Smith, the significant support of the Clinical Investigations Research Unit (Brighton and Sussex University Hospitals NHS Trust) and the editorial assistance of Robert Simonoff and A S Chamings. We would also like to acknowledge the dedication and support of Colin Barton from the Sussex & Kent ME/CFS Society and Janice Kent of ReMEmber.

## COPYRIGHT/LICENSE FOR PUBLICATION

The Corresponding Author has the right to grant on behalf of all authors and does grant on behalf of all authors, a worldwide licence to the Publishers and its licensees in perpetuity, in all forms, formats and media (whether known now or created in the future), to i) publish, reproduce, distribute, display and store the Contribution, ii) translate the Contribution into other languages, create adaptations, reprints, include within collections and create summaries, extracts and/or, abstracts of the Contribution, iii) create any other derivative work(s) based on the Contribution, iv) to exploit all subsidiary rights in the Contribution, v) the inclusion of electronic links from the Contribution to third party material where-ever it may be located; and, vi) licence any third party to do any or all of the above.

## ETHICS APPROVAL

Ethical approval was obtained from London-Brighton & Sussex Research Ethics Committee (REF:17/LO/0845).

## TRANSPARENCY

The manuscript’s guarantor affirms that the manuscript is an honest, accurate, and transparent account of the study being reported; that no important aspects of the study have been omitted; and that any discrepancies from the study as originally planned (and, if relevant, registered) have been explained.

